# Development of a Flexible Multiplex Urine RNA Assay for Detection and Differentiation of Kidney Allograft Injury

**DOI:** 10.1101/2025.08.25.25334374

**Authors:** Karen S. Keslar, Wenjie Xu, Anna A. Zmijewska, Dajana Margeta, Jonathan S. Bromberg, John J. Friedewald, Michael M. Abecassis, Kenneth A. Newell, Yvonne Morrison, Nancy D. Bridges, Peter S. Heeger, Roslyn B. Mannon, Robert L. Fairchild

**Author notes:** Correspondence: Dr. Roslyn B. Mannon, 983040 Nebraska Medical Center, Omaha, NE 68198-3040; Dr. Robert L. Fairchild, NB3-59, Cleveland Clinic, 9500 Euclid Avenue, Cleveland, OH 44195. Drs. Mannon and Fairchild are co-senior authors of this report.

## Abstract

Non-invasive approaches to detect kidney graft injury and distinguish donor-specific immune-mediated injury from other causes of inflammation are needed to guide recipient therapy while avoiding the morbidities of transplant biopsies. We used a multi-plex platform to interrogate RNA isolated from kidney transplant recipient urine to investigate gene expression patterns distinguishing grafts with no injury vs. ongoing injury and further differentiate acute T cell-mediated rejection (TCMR) from BK virus nephropathy (BKVN). As a training set we quantified expression of 796 immune function genes from 25 control recipients with stable graft function, 17 with biopsy-proven acute TCMR, and 13 with biopsy-proven BKVN. We identified a 20-gene signature that differentiated intragraft injury from grafts with stable function (area under the curve (AUC), 0.991) and a distinct 40-gene signature distinguishing acute TCMR from BKVN (AUC = 1.00). Validation in separate 118 urine RNA samples obtained at time of surveillance or for-cause biopsies from Clinical Trials in Organ Transplantation (CTOT)-08 and CTOT-19 studies showed AUC of 0.77 for the 20-gene injury signature and AUC of 0.79 for the 40-gene signature. Our results highlight the utility of this flexible, non-invasive biomarker platform for rapid detection and differentiation of immune processes causing ongoing kidney graft injury.

## 1 INTRODUCTION

The success of kidney transplantation to treat patients with end-stage kidney disease is compromised by the occurrence of T cell and antibody mediated rejection among other inflammatory mechanisms mediating allograft injury, all of which currently require a graft biopsy for diagnosis. Development and validation of non-invasive biomarkers capable of detecting ongoing allograft injury and differentiating the causes of the injury are needed. While transplant biopsies are safe, they are associated with defined risks and significant costs that could be reduced by accurate, noninvasive approaches. Moreover, intra-observer variation in pathology reads and tissue sample acquisition bias are known problems that complicate interpretation of biopsies (1–4), and both could be overcome by accurate non-invasive biomarkers.

The advent of molecular approaches to accurately detect increases in transcripts and proteins specific to tissues experiencing inflammation in cancer and autoimmune disease has been extended to kidney transplant patients. RNA from allograft biopsies has been interrogated by microarray analyses and molecular signatures identified for both immune and non-immune mediated injuries (5–8). While this “molecular microscope” and work by others to develop molecular phenotypes indicating acute and chronic graft injury has advanced detection (9–11), the approach is time consuming and requires the same invasive procedure to obtain graft tissue for RNA analysis as is the case for evaluation of graft histopathology.

Consequently, multiple efforts have focused on profiling RNA expression by microarray analyses of peripheral blood or isolated blood cell populations from kidney transplant patients. These include a gene expression profile (GEP) with a strong negative predictive value for the absence of subclinical kidney graft rejection and predictive of graft improvement following treatment for the subclinical rejection and a 17-GEP that detects subclinical rejection and stratifies the patients for risk of subsequent acute rejection and graft loss (12, 13). Importantly, these promising biomarkers have not been shown to differentiate types of kidney allograft injury beyond identifying rejection from the absence of rejection.

As graft-infiltrating immune cells and damaged urothelial cells are shed into the urine as a consequence of kidney graft inflammation, urinary RNA can be used to identify transcript markers of the injury (14). Work by others used a pre-amplification enhanced qPCR of urine RNA to identify transcripts associated with imminent or ongoing acute rejection as well as transcripts highly predictive of reversal of acute rejection and the risk of graft failure within six months of testing (15, 16). Other research groups identified urine RNA transcripts associated with kidney graft fibrosis and antibody mediated rejection (17). Despite these successes, qPCR approaches are hindered by the limited number of probes, the requirement for preamplification of target transcripts that can potentially lead to errors in detection, and the observation that the identified targets are sensitive for rejection but lack specificity for cause of graft injury (17–20).

In an effort to increase the range and strength of urine RNA transcript profiling to detect and differentiate subtypes of ongoing kidney graft injury we employed the NanoString nCounter platform as a multiplex approach to interrogate RNA isolated from the urine of kidney transplant patients. This approach provides quantitative transcript level data on up to 800 gene targets, does not include a pre-amplification step, and is amenable to detecting transcripts in samples with partially degraded RNA, which increases the efficacy of testing urine RNA. By exploiting the flexibility of using almost 800 distinct data points from each urine sample, we show that this strategy can be used to easily identify urinary GEP capable of detecting graft injury and distinguishing acute T cell mediated rejection (TCMR) from graft injury caused by BK virus nephropathy (BKVN).

## 2 MATERIALS AND METHODS

### 2.1 Kidney Transplant Patients

#### Training set samples

We procured 55 training set samples from renal transplant recipients enrolled in the Clinical Trials of Organ Transplantation (CTOT) studies CTOT-10 (21), CTOT-15 (22), and CTOT-16 (23), the Cleveland Clinic renal biorepository, and the University of Maryland. Clinical and demographic details of the training set subjects are listed in Table 1. All of the TCMR samples in the training set were collected in conjunction with indication biopsies. Control subjects were all enrolled in CTOT-15 or CTOT-16, which did not include protocol biopsies. Control subjects were defined as those with stable graft function (serum creatinine <2.0 mg/dl, no graft loss, treatment for acute rejection, or evidence of BK infection) through the entire follow-up period of the study (18 months for CTOT-15 and 12 months for CTOT-16). The training set consisted of: 25 urine samples from control subjects; 17 subjects with biopsy-proven acute TCMR of Banff grades I-III; and, 13 subjects with biopsy-proven BKVN.

**Table 1.**
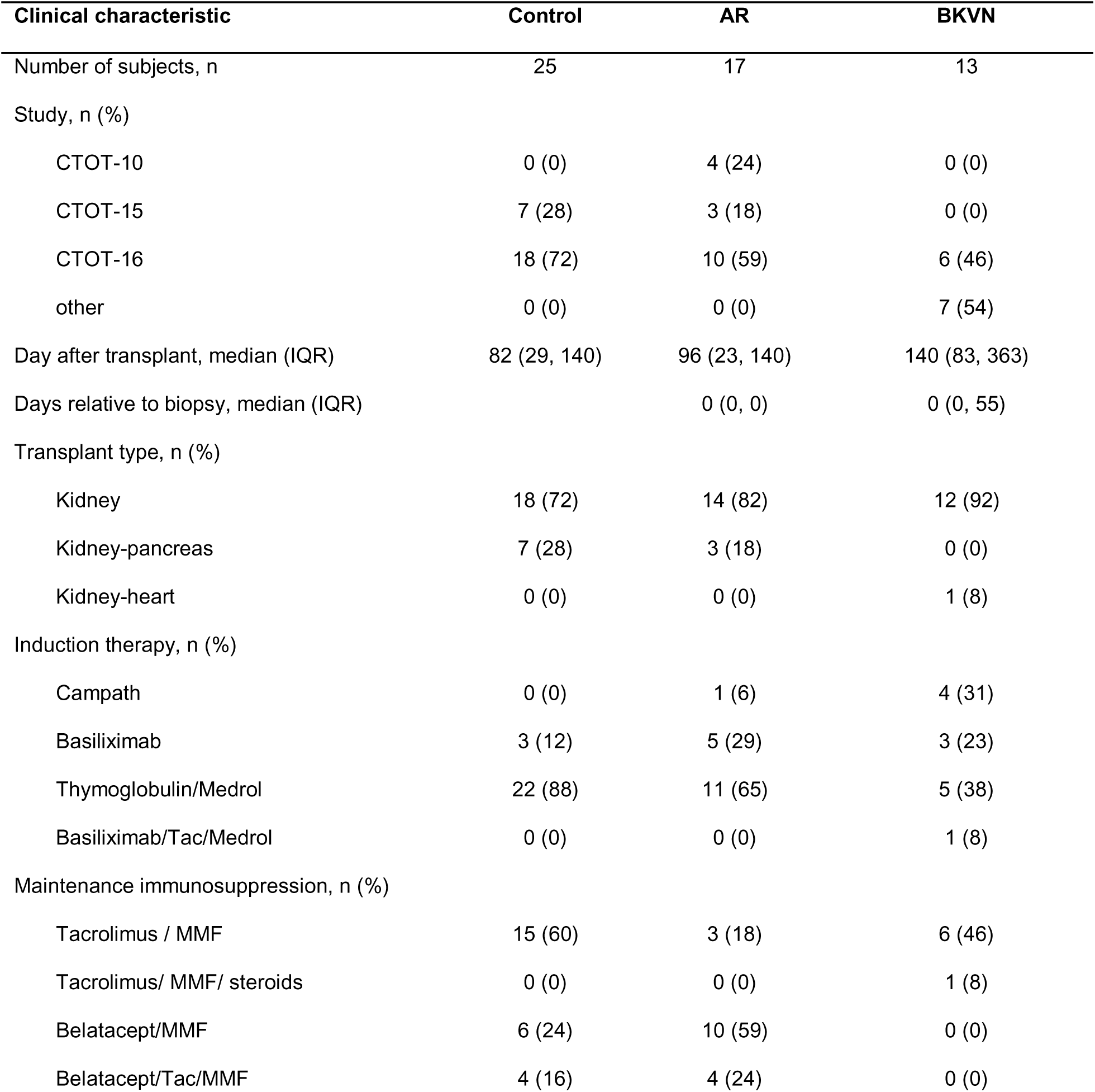
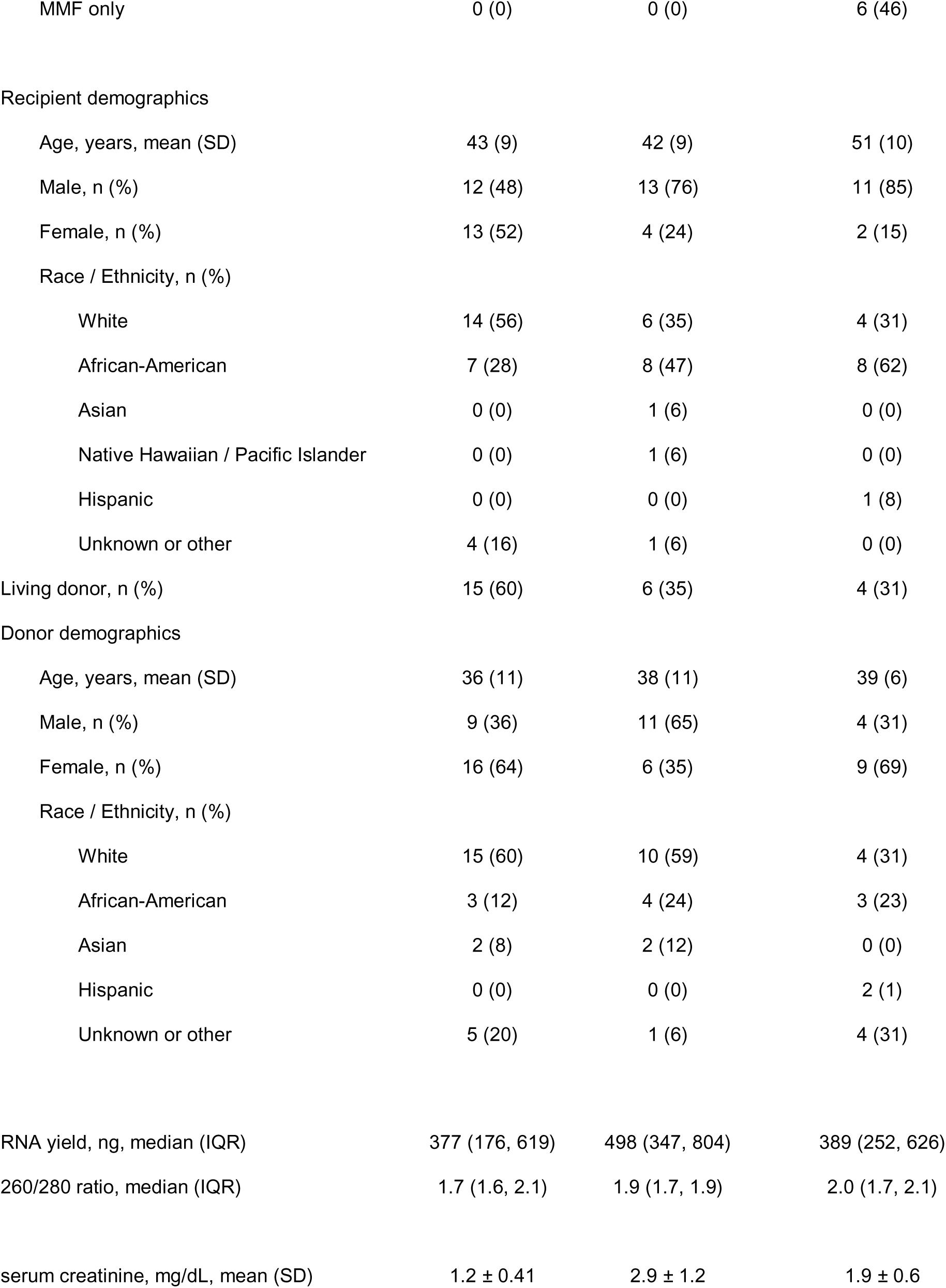
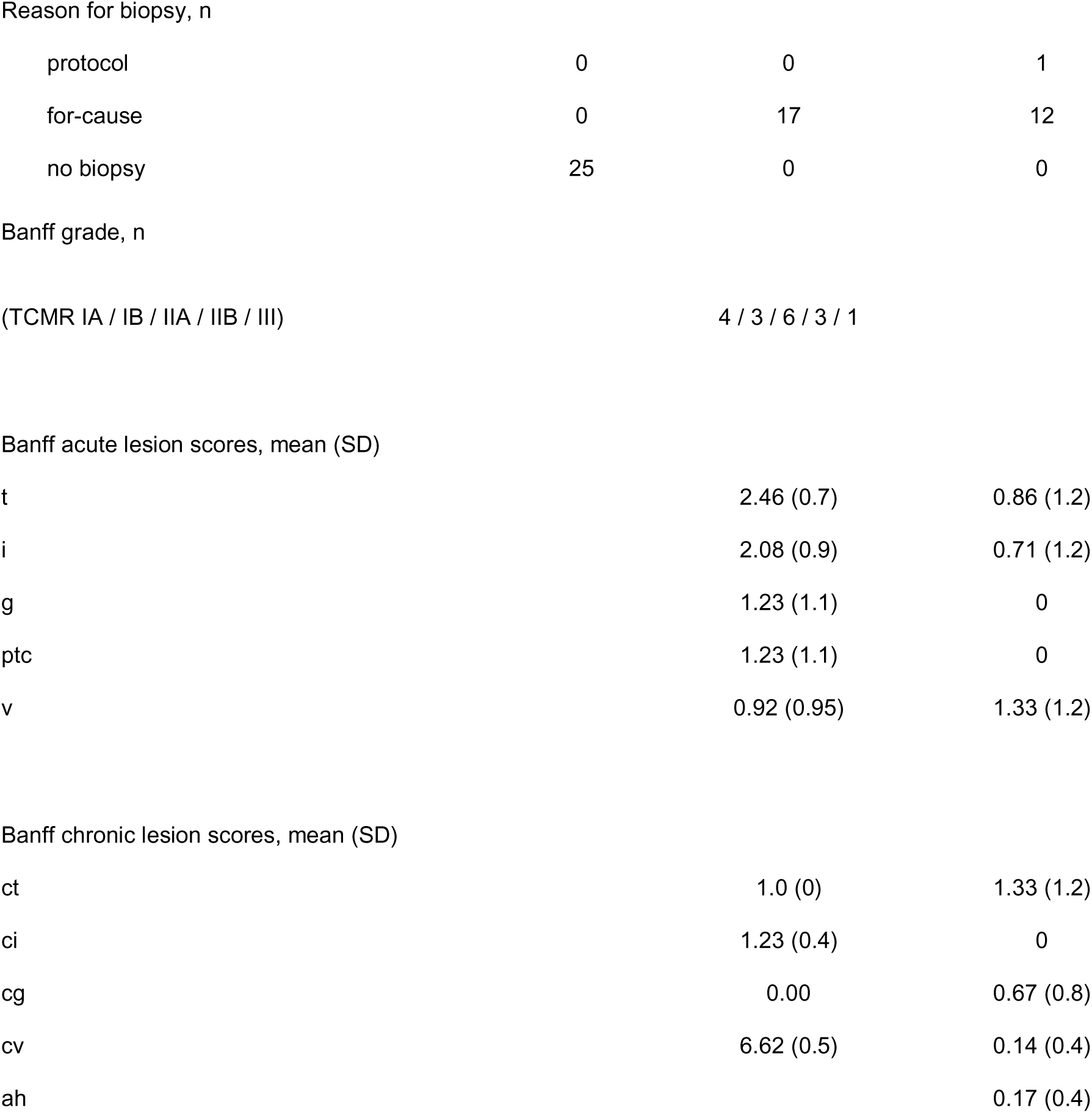
Clinical and demographic characteristics of the 55 training set subjects.

#### Validation set samples

We validated gene expression signatures using an independent set of blinded samples from 118 subjects enrolled in the CTOT-08 (12) and CTOT-19 studies. In CTOT-19, recipients of deceased donor kidneys with Kidney Donor Profile Index scores 20-90% were treated with ATG induction and randomized to receive a single intra-operative infusion of anti-TNFα monoclonal antibody (Infliximab/Remicade, Janssen, Beerse, Belgium) or saline. The half-life of Infliximab is < 2 weeks and all study samples were obtained after 6 months, making any effect of the peri-operative intervention an unlikely confounder of the study results. Maintenance immunosuppression consisted of tacrolimus, mycophenolate mofetil and prednisone. As the 2-year follow-up is not completed for all enrollees, the study remains blinded to the perioperative intervention. All subjects in these studies were biopsied at defined time points (surveillance biopsies) or to evaluate an acute change in allograft function (for-cause/clinically indicated biopsies). The clinical and demographic details of the test set subjects are described in Table 2. The validation set contained 100 samples from control subjects with normal biopsies and no indication of rejection of any type or BKVN, 12 from subjects with biopsy-proven TCMR, and 6 samples from subjects with BKVN. All of the study protocols were approved by the institutional review boards of the clinical sites where patients were enrolled and subjects provided informed consent. The clinical and research activities that we report here are consistent with the principles of the “Declaration of Istanbul on Organ Trafficking and Transplant Tourism” and the “World Medical Association Declaration of Helsinki on Ethical Principles for Medical Research Involving Human Subjects” (24, 25).

**Table 2.**
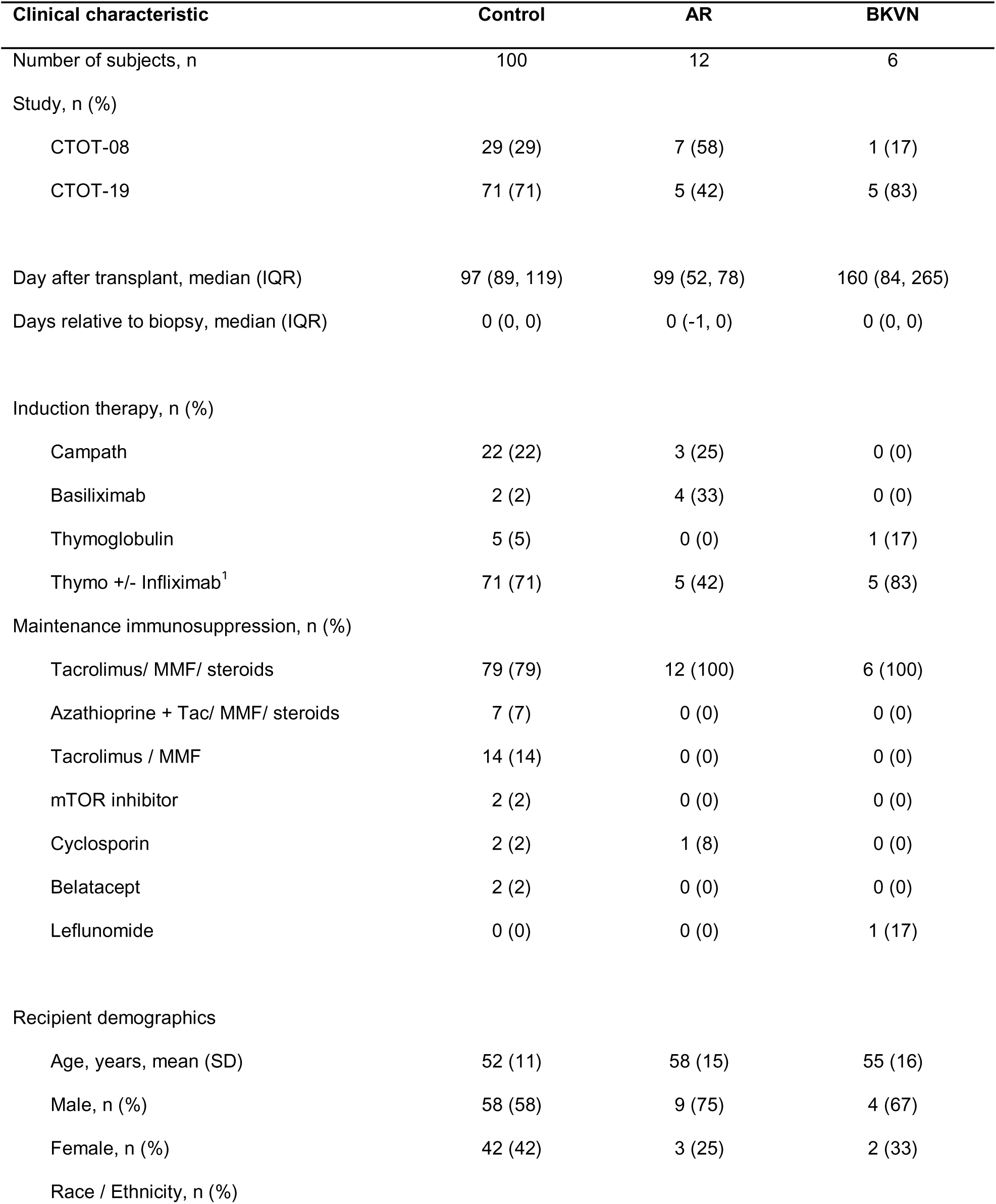
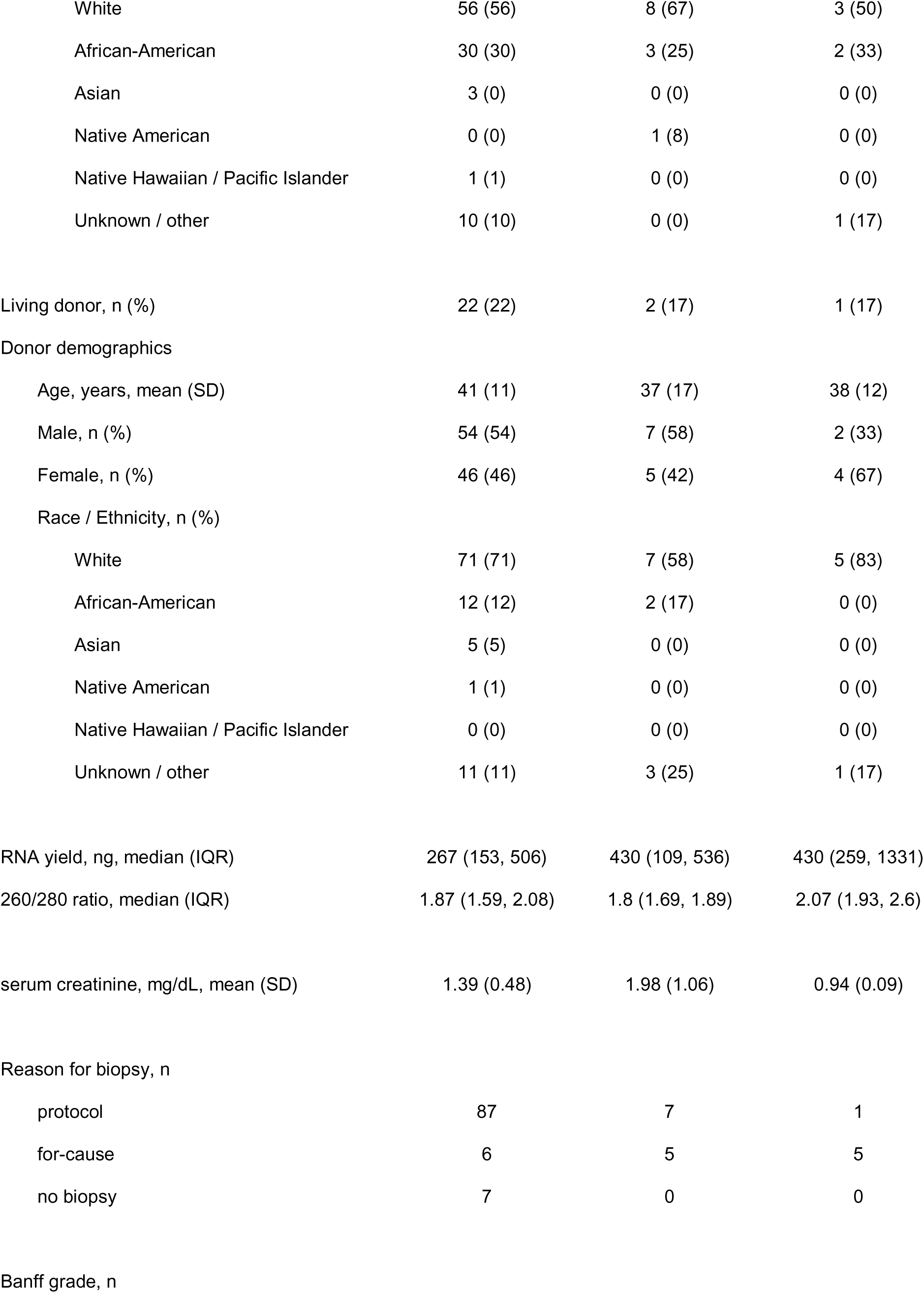
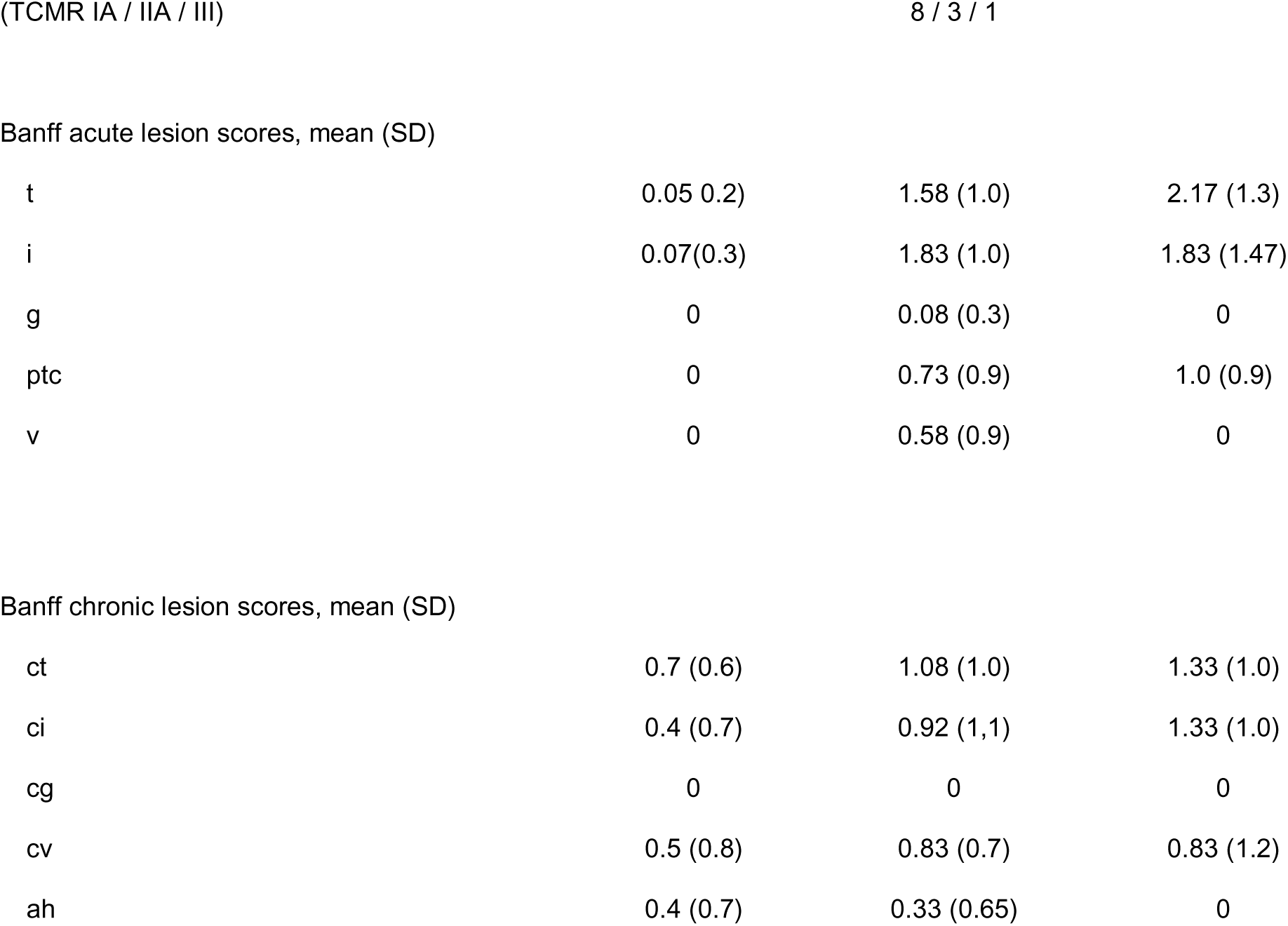
Clinical and demographic characteristics of the 118 validation set subjects. ^1^CTOT19 subjects were randomized into two groups– one group received infliximab and the other did not.

| **Clinical characteristic** | **Control** | **AR** | **BKVN** |
| --- | --- | --- | --- |
| Number of subjects, n | 100 | 12 | 6 |
| Study, n (%) |  |  |  |
| CTOT-08 | 29 (29) | 7 (58) | 1 (17) |
| CTOT-19 | 71 (71) | 5 (42) | 5 (83) |
| Day after transplant, median (IQR) | 97 (89, 119) | 99 (52, 78) | 160 (84, 265) |
| Days relative to biopsy, median (IQR) | 0 (0, 0) | 0 (-1, 0) | 0 (0, 0) |
| Induction therapy, n (%) |  |  |  |
| Campath | 22 (22) | 3 (25) | 0 (0) |
| Basiliximab | 2 (2) | 4 (33) | 0 (0) |
| Thymoglobulin | 5 (5) | 0 (0) | 1 (17) |
| Thymo +/- Infliximab<sup>1</sup> | 71 (71) | 5 (42) | 5 (83) |
| Maintenance immunosuppression, n (%) |  |  |  |
| Tacrolimus/ MMF/ steroids | 79 (79) | 12 (100) | 6 (100) |
| Azathioprine + Tac/ MMF/ steroids | 7 (7) | 0 (0) | 0 (0) |
| Tacrolimus / MMF | 14 (14) | 0 (0) | 0 (0) |
| mTOR inhibitor | 2 (2) | 0 (0) | 0 (0) |
| Cyclosporin | 2 (2) | 1 (8) | 0 (0) |
| Belatacept | 2 (2) | 0 (0) | 0 (0) |
| Leflunomide | 0 (0) | 0 (0) | 1 (17) |
| Recipient demographics |  |  |  |
| Age, years, mean (SD) | 52 (11) | 58 (15) | 55 (16) |
| Male, n (%) | 58 (58) | 9 (75) | 4 (67) |
| Female, n (%) | 42 (42) | 3 (25) | 2 (33) |
| Race / Ethnicity, n (%) |  |  |  |
| White | 56 (56) | 8 (67) | 3 (50) |
| African-American | 30 (30) | 3 (25) | 2 (33) |
| Asian | 3 (0) | 0 (0) | 0 (0) |
| Native American | 0 (0) | 1 (8) | 0 (0) |
| Native Hawaiian / Pacific Islander | 1 (1) | 0 (0) | 0 (0) |
| Unknown / other | 10 (10) | 0 (0) | 1 (17) |
| Living donor, n (%) | 22 (22) | 2 (17) | 1 (17) |
| Donor demographics |  |  |  |
| Age, years, mean (SD) | 41 (11) | 37 (17) | 38 (12) |
| Male, n (%) | 54 (54) | 7 (58) | 2 (33) |
| Female, n (%) | 46 (46) | 5 (42) | 4 (67) |
| Race / Ethnicity, n (%) |  |  |  |
| White | 71 (71) | 7 (58) | 5 (83) |
| African-American | 12 (12) | 2 (17) | 0 (0) |
| Asian | 5 (5) | 0 (0) | 0 (0) |
| Native American | 1 (1) | 0 (0) | 0 (0) |
| Native Hawaiian / Pacific Islander | 0 (0) | 0 (0) | 0 (0) |
| Unknown / other | 11 (11) | 3 (25) | 1 (17) |
| RNA yield, ng, median (IQR) | 267 (153, 506) | 430 (109, 536) | 430 (259, 1331) |
| 260/280 ratio, median (IQR) | 1.87 (1.59, 2.08) | 1.8 (1.69, 1.89) | 2.07 (1.93, 2.6) |
| serum creatinine, mg/dL, mean (SD) | 1.39 (0.48) | 1.98 (1.06) | 0.94 (0.09) |
| Reason for biopsy, n |  |  |  |
| protocol | 87 | 7 | 1 |
| for-cause | 6 | 5 | 5 |
| no biopsy | 7 | 0 | 0 |
| Banff grade, n |  |  |  |
| t | 0.05 (0.2) | 1.58 (1.0) | 2.17 (1.3) |
| i | 0.07 (0.3) | 1.83 (1.0) | 1.83 (1.47) |
| g | 0 | 0.08 (0.3) | 0 |
| ptc | 0 | 0.73 (0.9) | 1.0 (0.9) |
| v | 0 | 0.58 (0.9) | 0 |
| Banff chronic lesion scores, mean (SD) |  |  |  |
| ct | 0.7 (0.6) | 1.08 (1.0) | 1.33 (1.0) |
| ci | 0.4 (0.7) | 0.92 (1.1) | 1.33 (1.0) |
| cg | 0 | 0 | 0 |
| cv | 0.5 (0.8) | 0.83 (0.7) | 0.83 (1.2) |
| ah | 0.4 (0.7) | 0.33 (0.65) | 0 |

### 2.2 Isolation of urine RNA and NanoString analyses

Urine samples of 50-100 ml were collected within one week of for-cause or protocol scheduled surveillance biopsies, with 80% collected on the day of the biopsy with the times indicated in Tables 1 and 2. Samples were centrifuged at 2,000 x g for 30 minutes at 4°C. The pellets were washed with PBS, resuspended in RNAlater, stored at −80°C, and then shipped to the Cleveland Clinic. RNA was isolated from the urine sediment using Invitrogen PureLink RNA Mini Kit. Whenever possible, 100 ng of RNA was used for NanoString analysis. If the sample contained less than 100 ng of RNA, the entire sample was used. RNA was hybridized to a 796-gene Panel Plus codeset consisting of the NanoString PanCancer Immune Profiling gene panel with the addition of 26 genes that had been shown from published results or our own unpublished data to be involved in graft rejection or the development of fibrosis (Supplemental Table I).

The hybridized RNA samples were processed on the nCounter GEN2 Analysis System using the high sensitivity protocol and high-resolution data capture. Raw data were imported into nSolver4.0 (NanoString). Data normalization and gene expression analysis were performed using the nSolver Advanced Analysis plug-in version 2.0.115. Normalized log_2_ counts were used in all analyses. Differentially expressed genes (DEGs) were defined as those with log_2_ fold-change ≥1 or ≤ −1 and p ≤ 0.05.

### 2.3 Gene signatures

Statistical modeling and signature development from the gene expression levels in the training set samples were performed by NanoString Data Analysis Services (Seattle, WA). Data from a training set of 55 patient samples (17 TCMR, 13 BKVN and 25 C) was used to identify potential diagnostic signatures that can differentiate the three groups of patients. Based on differential expression profiles among the three groups, a two-stage method was used to develop signatures. First, a signature that could distinguish samples from grafts with “injury” (specifically TCMR or BKVN) from normal grafts (Control) was identified. Second, a signature that could distinguish TCMR from BKVN in the samples with injury was identified.

Raw Nanostring counts were normalized to the expression of ten housekeeping genes (CNOT10, COG7, ZC3H14, AGK, PPIA, EIF2B4, MTMR14, HPRT1, MRPS5, TUBB) that were chosen by the advanced analysis software using the Genorm algorithm, then log2 transformed to reduce the deviation from normality. Probes for RNA transcripts that had low overall counts or that showed minimal differential expression between any two groups (defined by ≤20 raw counts, or fold change between −1.3 and 1.3, or fold change p-value > 0.5) were removed, leaving 321 genes to be evaluated as predictors. Signature training was done using a machine learning algorithm based on the elastic net, a regularized regression method that linearly combines the L1 and L2 penalties of the lasso and ridge methods (26). Optimal tuning parameters were chosen by 10-fold cross-validation. The output from the machine learning algorithm is a set of predictor genes with corresponding weights that can be used to calculate a Linear Predictor Score, or LPS (LPS = ΣXiβi, where Xi = log_2_ expression level of gene i; {3i = assigned weight of gene i). Internal validation of each signature was done by 500 rounds of split-sample cross-validation.

Diagnostic cutoffs for both signatures were determined by optimizing the Youden index (27). The two signatures and their respective cutoffs were locked and then used to calculate LPS scores in a second, blinded validation set of urine RNA samples from 100 C subjects, 12 TCMR, and 6 BKVN. The raw NanoString counts for the genes in the two signatures were normalized and log_2_-transformed as described for the training set samples. For each validation set sample, the injury vs control linear predictor score was calculated, then for those samples that were classified as injured, the TCMR vs BKVN score was calculated.

### 2.4 Statistical analysis

ROC curve analysis, area under the curve plots, and volcano plots were performed in Sigmaplot 10.0; box and whisker plots and statistical analysis of variables between groups was performed in Graphpad Prism 9.0.

## 3 RESULTS

### 3.1 Urinary gene expression profile in T cell mediated acute rejection

We compared expression of 796 genes in RNA isolated from the urine of 25 kidney transplant recipients (KTRs) with stable graft function and no clinical indication of graft injury at the time of sample collection (and throughout the subsequent follow-up period for each study sample), but without a biopsy confirmation, to RNA isolated from 17 KTRs with graft dysfunction and biopsy-proven acute TCMR (Figure 1). Using a cutoff of log_2_ fold-change ≥ 1 or ≤ −1 and p ≤ 0.05 we observed a clear distinction between the two groups, with 41 up-regulated genes and 28 down-regulated genes in urine RNA from the TCMR group when compared with urine RNA from the control recipients with stable graft function. The up-regulated genes in urine samples from recipients with biopsy-proven TCMR included the chemokines CXCL9 and CXCL10, granzyme A, granzyme B, and perforin, which have been described in earlier studies of urine RNA using qPCR (28). These results confirm and validate those earlier findings and further demonstrate the robustness and reproducibility of GEP in urine sediment across KTR study populations and platforms.

**FIGURE 1.**
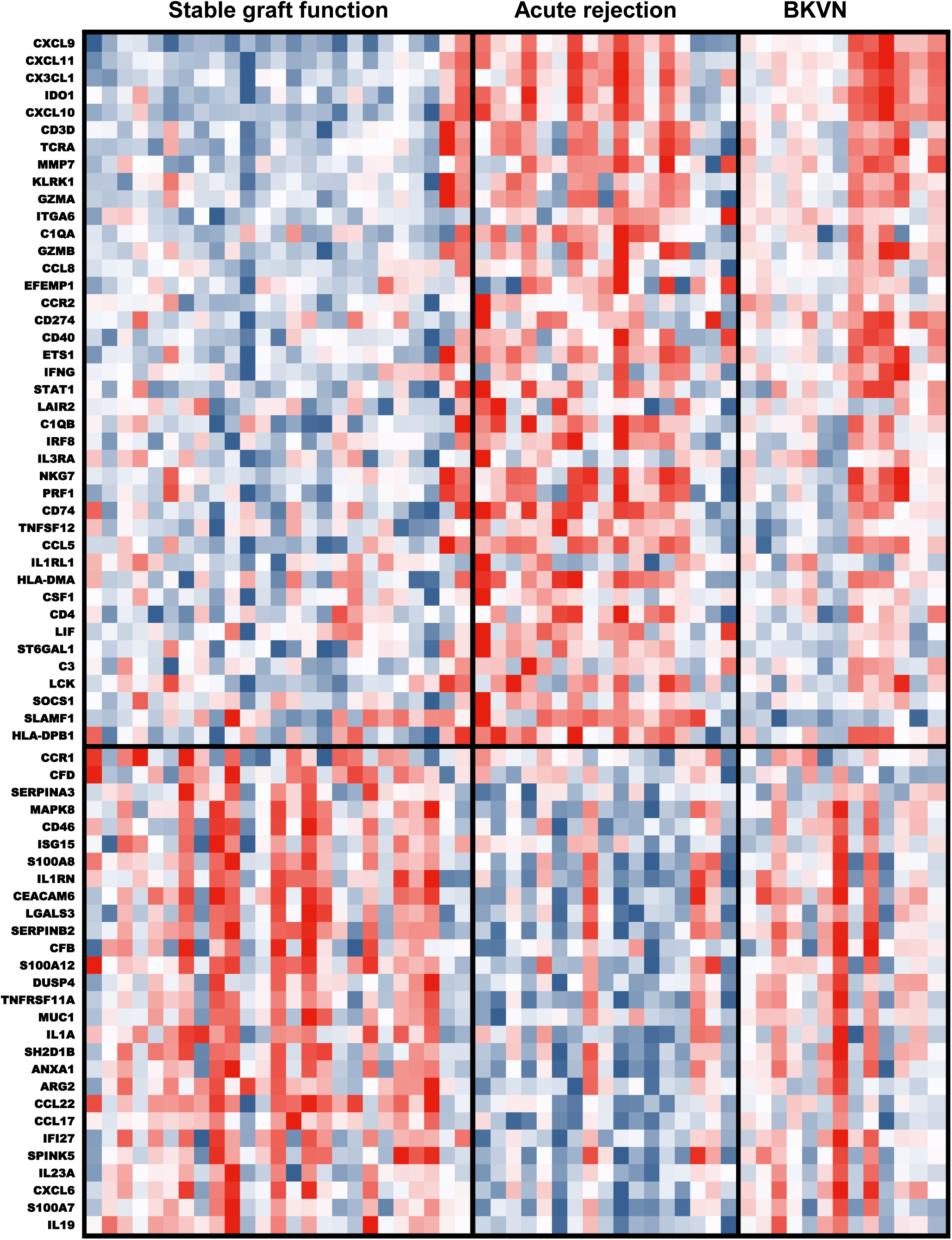
Up- and down-regulated gene expression patterns in urine RNA distinguish ongoing acute rejection of kidney grafts from grafts with stable function. The NanoString platform was used to measure expression levels of 796 mRNA transcripts in urine pellet RNA isolated from 25 kidney transplant recipients with stable graft function and no signs of acute rejection, 17 recipients with biopsy-proven T-cell mediated acute rejection, and 13 recipients with BK virus nephropathy diagnosed by biopsy. Using a cutoff of log_2_ fold-change ≥1 or ≤ −1 and p ≤ 0.05 to define differentially expressed genes (DEGs), 41 genes were up-regulated and 28 down-regulated during ongoing acute rejection when compared to control, with red in the heat map indicating transcript expression greater than the mean expression level for each given gene in all samples tested and blue indicating expression lower than the mean.

The GEP did not apply to all study subjects: urine RNA from two recipients with stable graft function showed the up-and down-regulation gene expression observed in the TCMR profile and urine samples from two recipients with biopsy indicated TCMR showed patterns that did not differentiate them from the controls. Overall, these initial studies indicated that this multiplex detection of differentially expressed genes (DEG) has the potential to provide useful diagnostic information to differentiate acute TCMR from the absence of inflammation.

### 3.2 Gene expression profiles shared and distinguishing acute TCMR and BKVN in the training set

Prior studies of GEP have indicated overlap of up-regulated genes observed in recipient urine during acute TCMR with other inflammatory conditions including both bacterial and BK virus infections (29). To address this potential overlap, we compared urinary GEP of 13 BKVN^+^ KTRs to those 25 KTRs with stable graft function using the 796 gene code set and observed 33 up-regulated and 30 down-regulated genes in KTRs with BKVN. There was clear up-regulation of many of the proinflammatory genes in urine of BKVN recipients as observed during acute rejection; however, the down-regulated gene expression observed during acute rejection was not observed in urine RNA from the recipients with BKVN and this expression looked similar to the up-regulated genes expressed in urine from the control recipients (Figure 1). As anticipated we observed significant overlap between DEGs of BKVN and TCMR with a total of 21 up-regulated and 3 down-regulated genes shared by both conditions (Figure 2 and Table 3). The shared genes included those with the highest fold-changes in acute rejection (CXCL9, CXCL10, TCRA, GZMA, GZMB, IFNG), transcripts encoding chemoattractants and functions of effector T cells. Further analysis showed enrichment of 20 up-regulated and 25 down-regulated genes specific to patients with acute TCMR and 12 up-regulated and 27 down-regulated genes specific to patients with BKVN (Figure 3 and Table 3).

**FIGURE 2.**
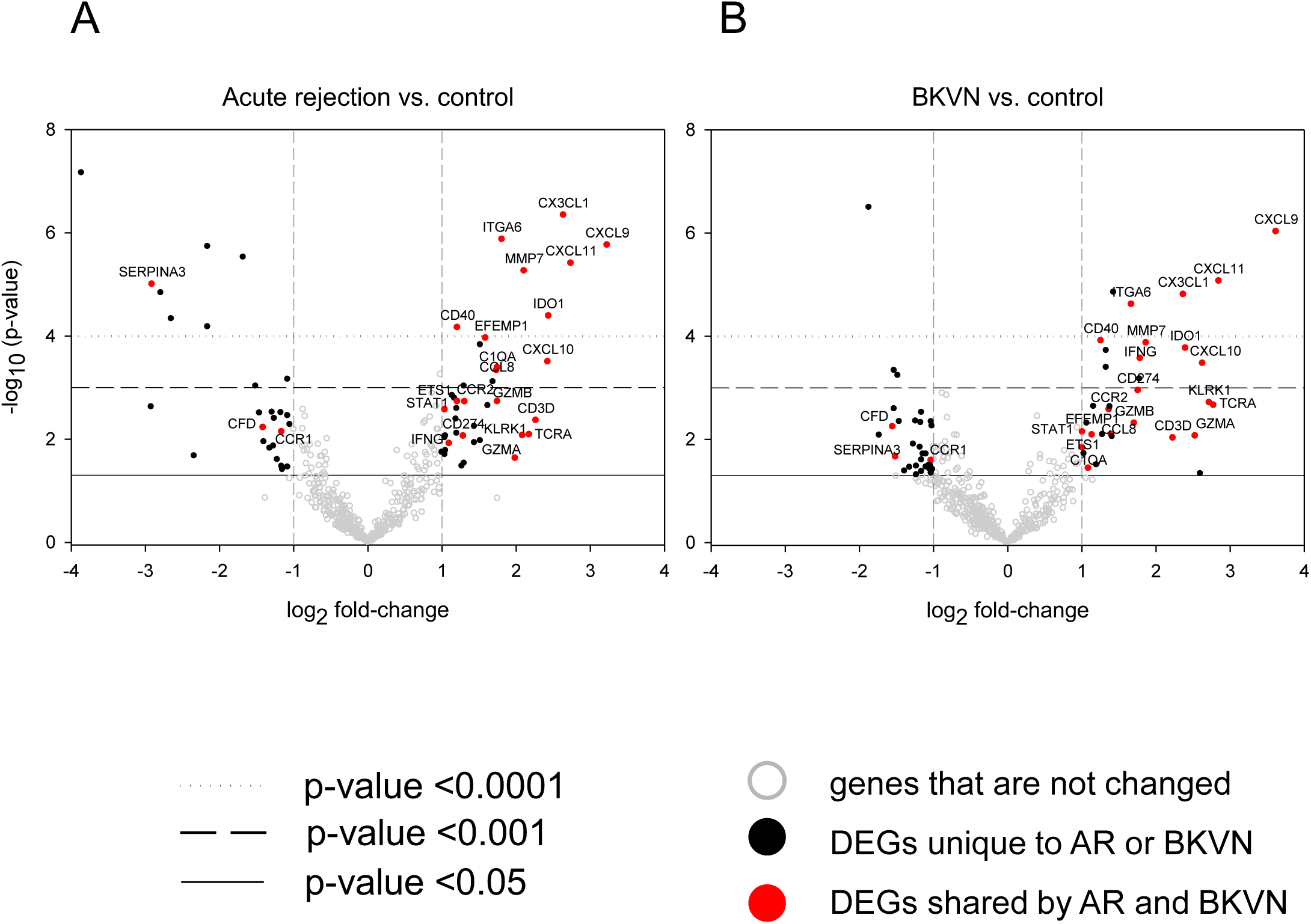
Shared gene expression changes in urine RNA of KTR with TCMR or BKVN. Volcano plots illustrate differences in expression of 796 genes in urine RNA isolated from the 55 training set samples. Log2 fold-change is shown on the X axis and −log10 p-value is on the Y axis A) Gene expression changes in TCMR (n = 17) and C (n = 25). The DEGs that are only in TCMR vs C are indicated by solid black circles. B) gene expression changes in BKVN (n = 13) and C (n = 25). DEGs that are only in BKVN vs C are indicated by solid black circles. In both panels, genes that fall below the cutoffs for differential expression (log_2_ fold-change ≥ 1 or ≤ −1 and p ≤ 0.05) are indicated by open gray circles. DEGs that are found in both TCMR vs C AND BKVN vs C are represented by red circles and the names are listed on the plot. Twenty one genes are upregulated and 3 downregulated in TCMR and BKVN compared to control. Raw p-values calculated by a two-tailed t test are indicated by horizontal lines as shown in the legend. The DEGs and whether they are common to both TCMR and BKVN or unique are listed in Table 3.

**FIGURE 3.**
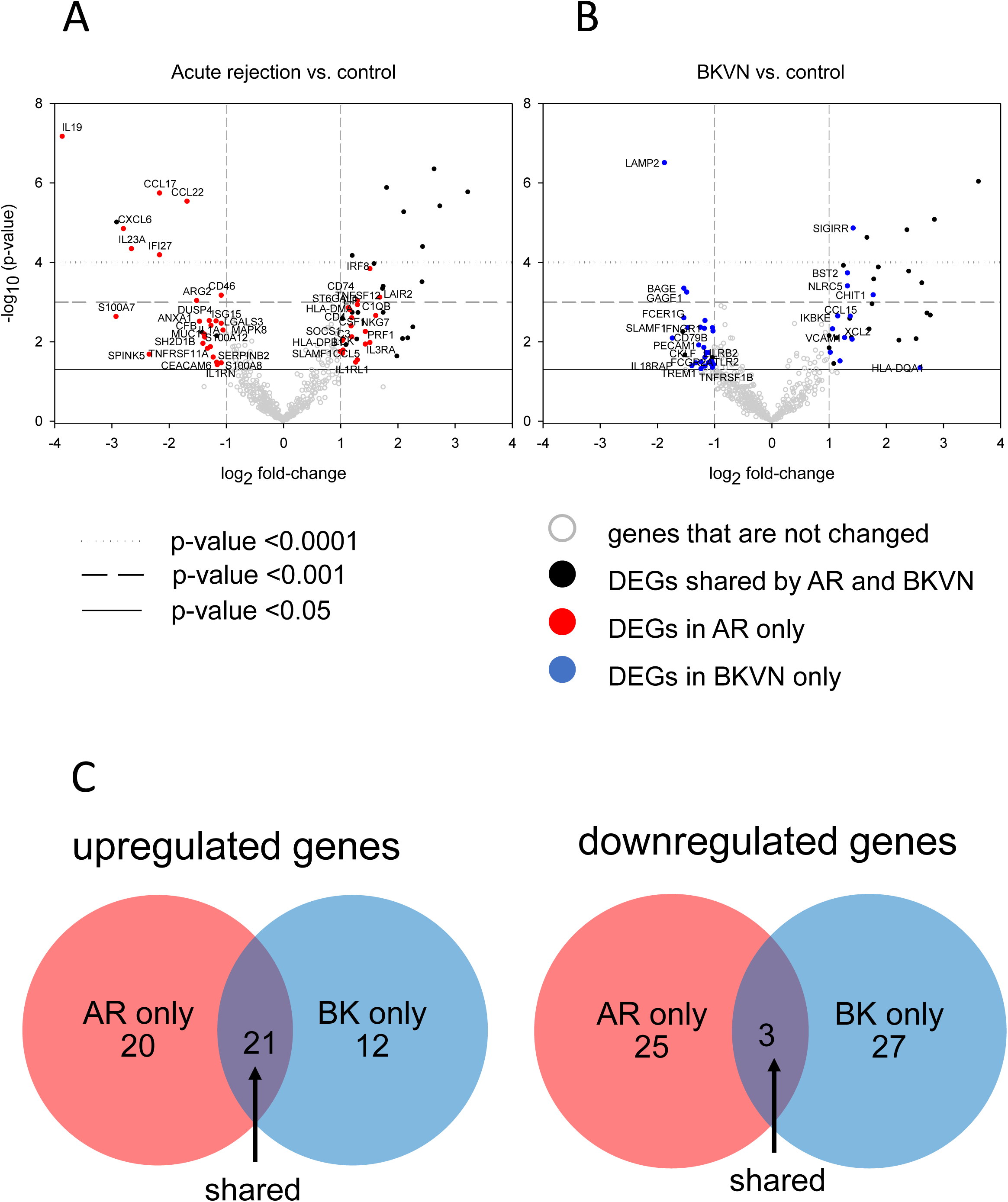
Urinary DEGs that differ between TCMR and BKVN. Volcano plots illustrate differences in expression of 796 genes in urine RNA isolated from the 55 training set samples. Log2 fold-change is shown on the X axis and −log10 p-value is on the Y axis. A) Gene expression changes in TCMR (n = 17) and C (n = 25). The DEGs that are only in TCMR vs C are indicated by solid red circles and are labeled on the plot. B) gene expression changes in BKVN (n = 13) and C (n = 25). DEGs that are only in BKVN vs C are indicated by solid blue circles and are labeled. In both panels, genes that fall below the cutoffs for differential expression (log_2_ fold-change ≥ 1 or ≤ −1 and p ≤ 0.05) are indicated by open gray circles. DEGs that are found in both TCMR vs C AND BKVN vs C are represented by solid black circles. Numbers of distinct and shared DEGs that are up- and down-regulated vs. controls are shown in the Venn diagrams (panel C) and are listed in Table 3.

**Table 3.** Unique and shared differently expressed genes in TCMR and BKVN.

**DEGs in TCMR only**
| **Gene** | **Direction** | **Gene** | **Direction** |
| --- | --- | --- | --- |
| LAIR2 | up | S100A8 | down |
| C1QB | up | IL1RN | down |
| IRF8 | up | CEACAM6 | down |
| IL3RA | up | LGALS3 | down |
| NKG7 | up | SERPINB2 | down |
| PRF1 | up | CFB | down |
| CD74 | up | S100A12 | down |
| TNFSF12 | up | DUSP4 | down |
| CCL5 | up | TNFRSF11A | down |
| IL1RL1 | up | MUC1 | down |
| HLA-DMA | up | IL1A | down |
| CSF1 | up | SH2D1B | down |
| CD4 | up | ANXA1 | down |
| LIF | up | TCMRG2 | down |
| ST6GAL1 | up | CCL22 | down |
| C3 | up | CCL17 | down |
| LCK | up | IFI27 | down |
| SOCS1 | up | SPINK5 | down |
| HLA-DPB1 | up | IL23A | down |
| MAPK8 | down | CXCL6 | down |
| CD46 | down | S100A7 | down |
| ISG15 | down | IL19 | down |

DEGs in BKNV only
| Gene | Direction | Gene | Direction |
| --- | --- | --- | --- |
| HLA-DQA1 | up | FCGR2B | down |
| CHIT1 | up | PECAM1 | down |
| SIGIRR | up | TGFB1 | down |
| XCL2 | up | CKLF | down |
| CCL15 | up | TLR4 | down |
| BST2 | up | IFNGR1 | down |
| NLRC5 | up | CCRL2 | down |
| VCAM1 | up | FCGR2A | down |
| CT45A1 | up | TREM1 | down |
| IKBKE | up | CXCL16 | down |
| NFATC2 | up | ADORA2A | down |
| ULBP2 | up | IFITM2 | down |
| CD14 | down | IL18RAP | down |
| CD79B | down | LILRB2 | down |
| PRKCD | down | GAGE1 | down |
| TNFRSF1B | down | BAGE | down |
| CD36 | down | FCER1G | down |
| HCK | down | SELL | down |
| TLR2 | down | LAMP2 | down |

DEGS common to TCMR and BK
| Gene | Direction | Gene | Direction |
| --- | --- | --- | --- |
| CXCL9 | up | ITGA6 | up |
| CXCL11 | up | CCL8 | up |
| TCRA | up | CCR2 | up |
| KLRK1 | up | CD40 | up |
| CXCL10 | up | EFEMP1 | up |
| GZMA | up | C1QA | up |
| IDO1 | up | STAT1 | up |
| CX3CL1 | up | ETS1 | up |
| CD3D | up | CCR1 | down |
| MMP7 | up | SERPINA3 | down |
| IFNG | up | CFD | down |
| CD274 | up | SLAMF1 | up-TCMR, |
| GZMB | up |  | down-BKVN |

### 3.3 Developing signatures to detect graft injury and distinguish TCMR

We used a two-step process to develop our signatures. In both steps, normalized log_2_-transformed Nanostring counts of 321 transcripts measured in the urine RNA of the training set subjects were used as predictors in a machine learning algorithm based on the elastic net. First, we looked for a combination of transcript levels that could distinguish urine from grafts with injury (TCMR or BKVN) from stable grafts with good function (C) The analysis yielded a 20-gene model:

LPS _INJ v C_ = (−0.199 x ATF1) + (−0.105 x CCL17) + (−0.004 x CCL22) + (0.011 x CCL3) + (0.135 x CD274) + (0.010 x CX3CL1) + (0.091 x CXCL10) + (0.189 x CXCL11) + (0.131 x CXCL9) + (−0.309 x DHX16) + (−0.038 x ELK1) + (−0.017 x GPATCH3) + (−0.102 x IFIT1) + (−0.139 x IL19) + (0.513 x ITGA6) + (−0.478 x MERTK) + (0.100 x SF3A3) + (0.136 x SIGIRR) + (0.009 x TNFRSF11B) + (−0.401 x TNFSF4).

ROC curve analysis indicated this signature has an AUC of 0.99 (95% CI 0.93 – 1.00; p < 0.0001). Using the diagnostic cutoff of −2.68, this signature has 97% sensitivity and 100% specificity to distinguish grafts with TCMR or BKVN from those without. The generalizability of the model to other datasets was determined by 500 rounds of split-sample cross-validation, which yielded a mean cross-validated AUC of 0.84 with a 95% confidence interval of 0.68 – 0.96 (Figure 4).

**FIGURE 4.**
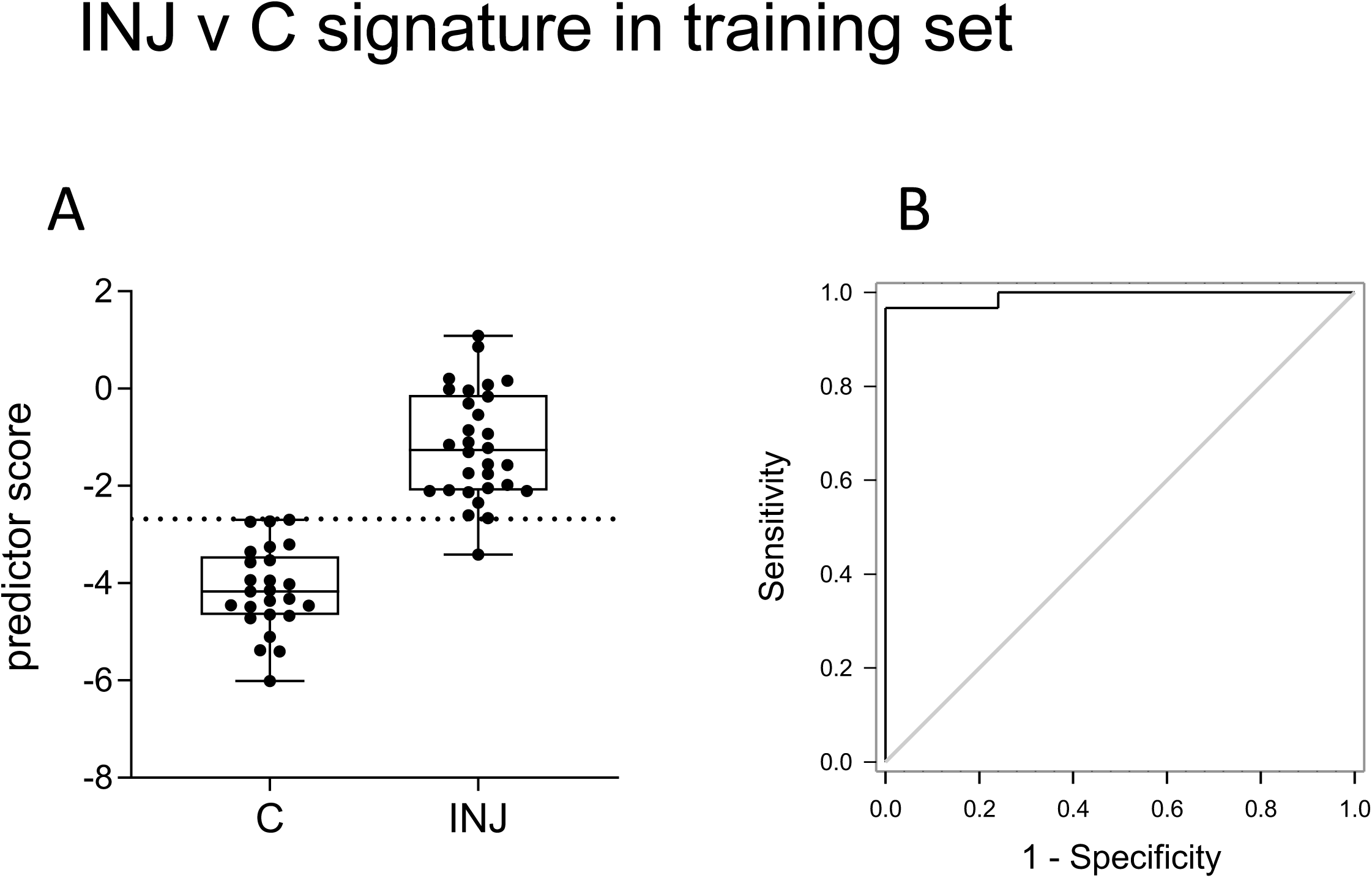
A 20-gene signature in urine RNA distinguishes control kidney grafts from those with TCMR or BKVN. Elastic net regression was used to determine a linear predictor score based on the weighted expression levels of 20 genes in urine RNA from training set samples. A) Box and whisker plots of the linear predictor scores in samples from 25 KTR with stable grafts (C) and 30 with either TCMR or BKVN (INJ). B) ROC curve analysis of the signature yielded an AUC = 0.99 (95% CI 0.93 – 1.00; p < 0.0001). With a diagnostic cutoff of −2.68 (indicated by a dotted line on the box plot) the sensitivity is 0.97 and specificity is 1. 500 rounds of split-sample cross-validation yielded a mean cross-validated AUC of 0.84 with a 95% confidence interval of 0.68 – 0.96. The 20 genes and their weights are listed in Table 4.

**Table 4.** Signature genes and weights. Using elastic net multinomial regression we developed a two-stage algorithm to first discriminate well-functioning grafts from those with injury based on expression of 20 genes in urine RNA, then to determine whether the injury is the result of AR or BK based on the expression levels of 40 genes. The genes and their regression coefficients are listed.

| 20 gene injury vs<br>stable |  | 40 gene TCMR vs BKVN |  |  |  |
| --- | --- | --- | --- | --- | --- |
| Gene | Weight | Gene | Weight | Gene | Weight |
| ATF1 | -0.19861 | ATF1 | 0.67259 | ITGB1 | 0.242935 |
| CCL17 | -0.10518 | BATF | -0.27108 | LAMP2 | 0.375836 |
| CCL22 | -0.00389 | BST2 | -0.42878 | LIF | 0.036167 |
| CCL3 | 0.010096 | C2 | 0.17646 | MAF | -0.15088 |
| CD274 | 0.135503 | CCL17 | -0.07676 | MAPK9 | -0.69094 |
| CX3CL1 | 0.010487 | CD274 | -0.19171 | MERTK | 0.087561 |
| CXCL10 | 0.091161 | CD68 | 0.1579 | NFATC1 | 0.044157 |
| CXCL11 | 0.189903 | CD86 | 0.29252 | NLRC5 | -0.03255 |
| CXCL9 | 0.131362 | CD99 | 0.14595 | PBEF1 | -0.03148 |
| DHX16 | -0.30984 | CEACAM8 | 0.01713 | PIN1 | 1.007383 |
| ELK1 | -0.03804 | EGFR | -0.12748 | PRAME | 0.124029 |
| GPATCH3 | -0.01662 | ELK1 | 0.2168 | PRPF38A | -0.23596 |
| IFIT1 | -0.10162 | GBP1 | -0.0733 | SERPINA3 | -0.06524 |
| IL19 | -0.13982 | GPATCH3 | 0.49989 | SLAMF1 | 0.159386 |
| ITGA6 | 0.512774 | GPI | 0.87842 | SPA17 | -0.63723 |
| MERTK | -0.47803 | GTF3C1 | -0.45258 | ST6GAL1 | 0.149539 |
| SF3A3 | 0.10038 | HLA-DRB3 | 0.09013 | STAT4 | 0.19475 |
| SIGIRR | 0.135693 | HMGB1 | -0.72851 | TNF | 0.488925 |
| TNFRSF11B | 0.008826 | IL21R | 0.46897 | TNFRSF11B | -0.60182 |
| TNFSF4 | -0.40289 | IL6ST | 0.39396 | YTHDF2 | 1.379289 |

In the second step we analyzed only the training set samples from the TCMR and BKVN groups to develop a signature to distinguish the two kidney graft pathologies. The 40-gene model, shown below, had an AUC of 1.0 (95% CI 0.94 – 1.0; p < 0.001), and 100% sensitivity and specificity when using a cutoff of 22.73. After 500 rounds of cross-validation, the mean estimated AUC was 0.95, 95% CI 0.78 – 1. This is the expected value of the AUC in an independent set of samples not used to develop the signature (Figure 5).

LPS _TCMR vs BKVN_ = (0.672 x ATF1) + (−0.271 x BATF) + (−0.429 x BST2) + (0.176 x C2) + (−0.077 x CCL17) + (−0.192 x CD274) + (0.158 x CD68) + (0.292 x CD86) + (0.146 x CD99) + (0.017 x CEACAM8) + (−0.127 x EGFR) + (0.217 x ELK1) + (−0.073 x GBP1) + (0.499 x GPATCH3) + (0.878 x GPI) + (−0.452 x GTF3C1) + (0.09 x HLA-DRB3) + (−0.728 x HMGB1) + (0.469 x IL21R) + (0.394 x IL6ST) + (0.243 x ITGB1) + (0.376 x LAMP2) + (0.036 x LIF) + (−0.151 x MAF) + (−0.691 x MAPK9) + (0.088 x MERTK) + (0.044 x NFATC1) + (−0.032 x NLRC5) + (−0.031 x PBEF1) + (1.007 x PIN1) + (0.124 x PRAME) + (−0.236 x PRPF38A) + (−0.065 x SERPINA3) + (0.159 x SLAMF1) + (−0.637 x SPA17) + (0.149 x ST6GAL1) + (0.195 x STAT4) + (0.489 x TNF) + (−0.602 x TNFRSF11B) + (1.379 x YTHDF2)

**FIGURE 5.**
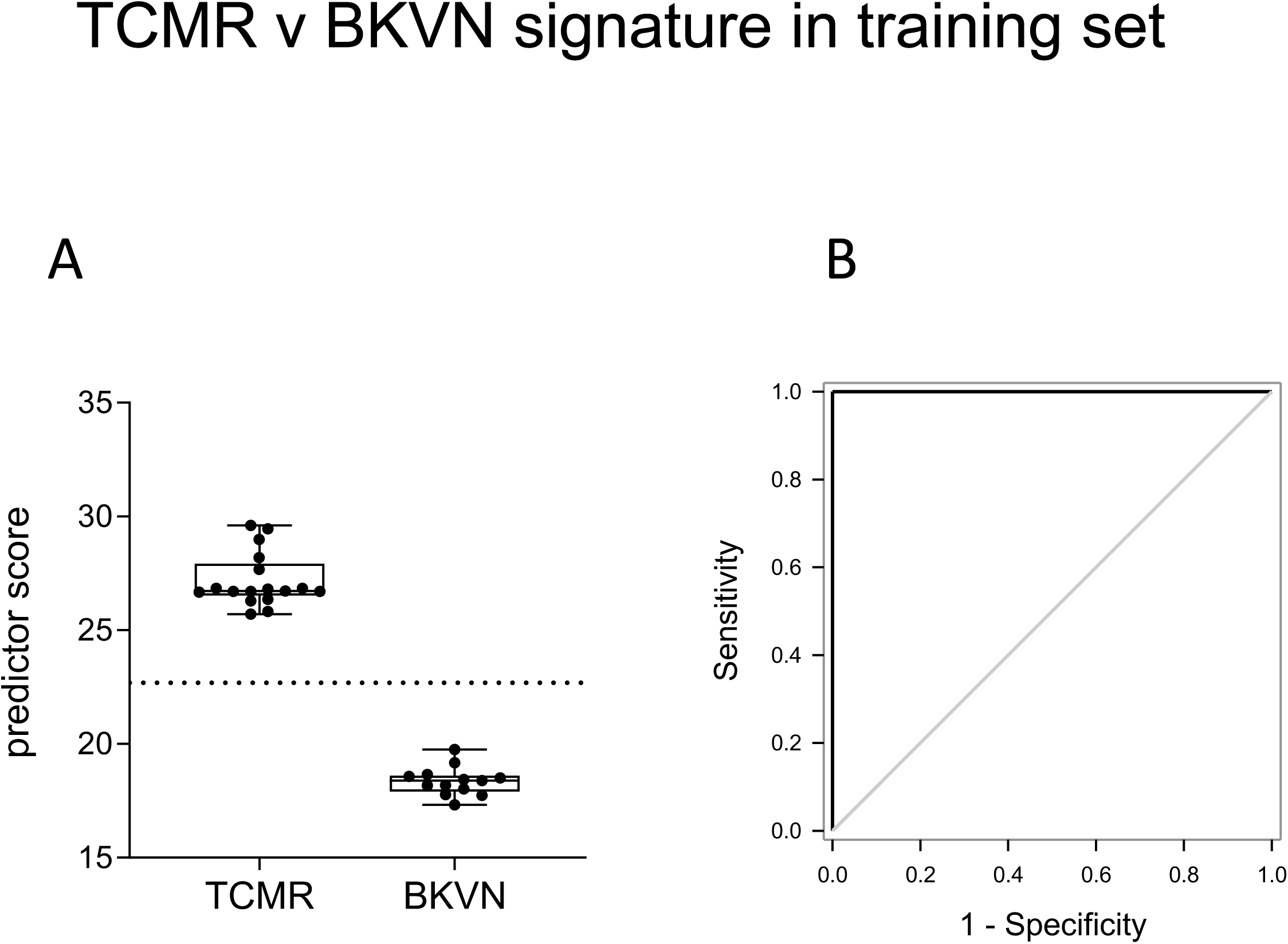
A 40-gene signature in urine RNA distinguishes injured grafts with TCMR from those with BKVN. Elastic net regression was used to determine a linear predictor score based on the weighted expression levels of 40 genes in urine RNA from training set samples. (A) Box and whisker plots of linear predictor scores in 17 TCMR and 13 BKVN samples. (B) ROC curve analysis yielded an AUC of 1.0 (95% CI 0.942 – 1.00; p < 0.001). Using the diagnostic cutoff of 22.73 (indicated by a dotted line on the box plot) the sensitivity, specificity, negative predictive value, and positive predictive value were all 1. The mean estimated AUC was from 500 rounds of split-sample cross-validation was 0.952, 95% CI 0.78 – 1. The 40 genes and their regression coefficients are listed in Table 4.

As a validation of the signatures to first distinguish graft injury vs. stability and then distinguish acute TCMR from BKVN, we applied the models to an independent set of 118 urine RNA samples obtained from 118 kidney transplant recipients at the time of a surveillance or for-cause biopsy (Figure 6). These analyses revealed an AUC for the 20-gene signature to detect graft injury of 0.77, 95% CI 0.64-0.89, p = 0.0003) and an AUC for the 40-gene signature to distinguish acute rejection vs. BKVN of 0.79, 95% CI 0.56-1.0, p < 0.05).

**FIGURE 6.**
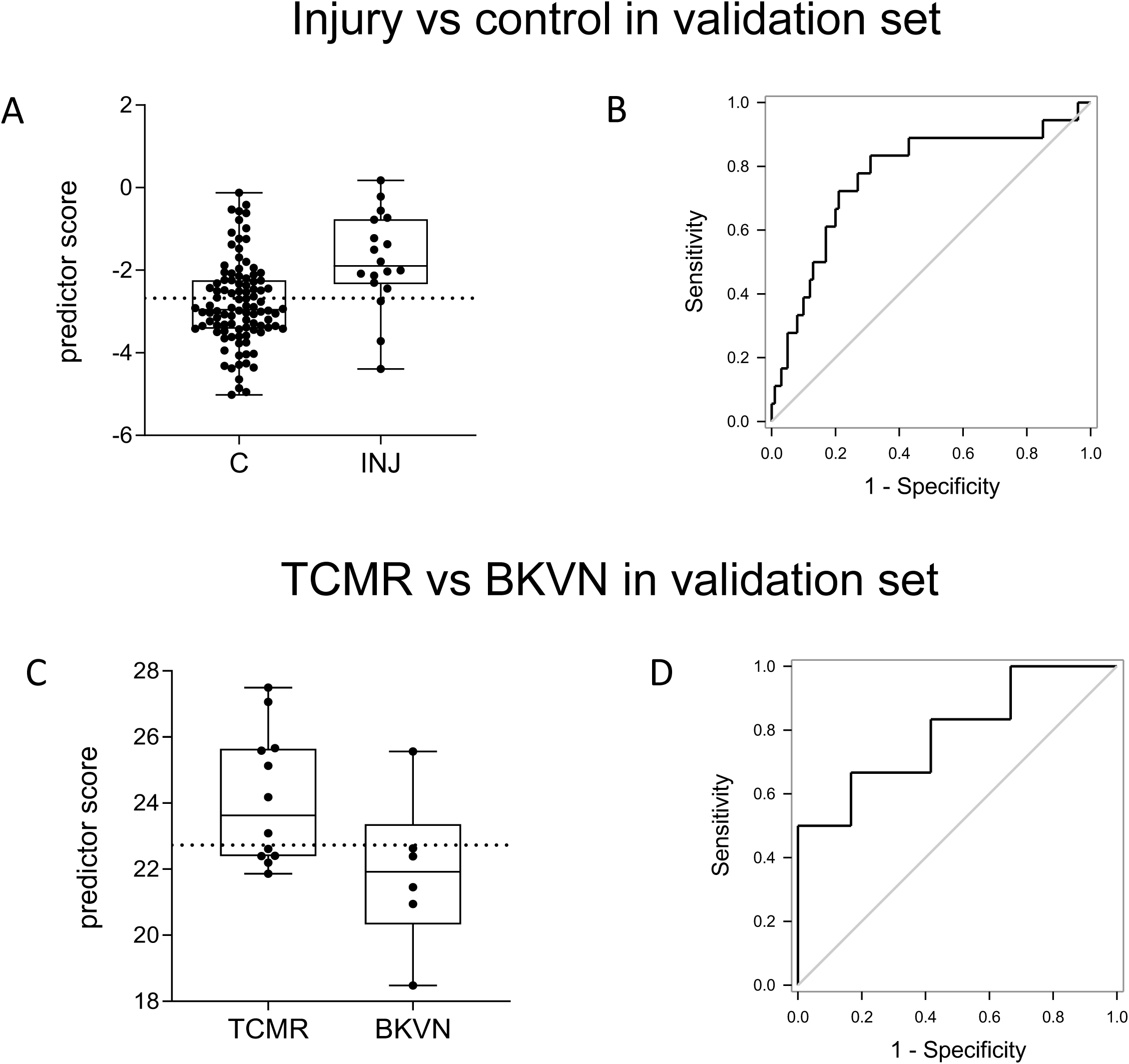
Validation of gene signatures in a separate cohort of KTR urine RNA samples. The 20- and 40-gene classifiers were used to calculate signature scores in a validation set of 118 urine samples collected at the time of biopsy (100 C, 12 TCMR, and 6 BKVN). A) Box and whisker plots of the 20-gene injury vs control linear predictor scores for 100 C and 18 TCMR or BKVN (INJ) samples. The diagnostic cutoff of −2.68 determined in the training set is indicated by a dotted line. B) ROC curve. The injury vs control signature had an AUC of 0.77, 95% CI 0.64-0.89, p = 0.0003 in the validation set. Sensitivity and specificity were 0.83 and 0.69, respectively, with Negative Predictive Value of 0.81 and Positive Predictive Value of 0.73. C) Box and whisker plots of the 40-gene TCMR vs BKVN linear predictor scores for 12 TCMR and 6 BKVN samples. The diagnostic cutoff of 22.73 determined in the training set samples is indicated by a dotted line. D) ROC curve. The TCMR vs BKVN signature had an AUC of 0.79, 95% CI 0.56-1, p < 0.05 in the validation samples. Sensitivity and specificity were 0.67 and 0.0.83, respectively, with Negative Predictive Value of 0.71 and Positive Predictive Value of 0.80.

Within the validation set the 20-gene signature correctly identified 59% of grafts having biopsy diagnosed no injury/no evidence of rejection. (Table 5) However, 41 samples (41%) from patients with graft biopsy diagnosed absence of rejection had a urine RNA 20-gene signature score indicating injury. Sixteen of these 41 patients had a urine RNA 40-gene signature score indicating acute TCMR and the other 25 patients had a score indicating BKVN (Table 5). Of the 12 urine RNA samples obtained from KTRs at the time of biopsy-proven rejection by the Central Pathology Core, the gene signatures correctly identified 50%. 6 AR were called AR; 2 AR were called C; 4 AR were called BK. While the NanoString profile of 2 samples showed the no injury signature, local pathology reads showed no rejection (discordant from the central reads) suggesting that the biomarker diagnosis is potentially correct. From the 6 patients with biopsy-diagnosed BKVN, the signatures correctly identified 4, with one sample incorrectly classified as an acute rejection and the other as having no injury.

**Table 5.** Summary of gene signature performance in validation samples. We measured gene expression levels in the urine RNA from a separate set of 118 blinded samples collected as part of the CTOT-08 and CTOT-19 studies. Using the log2-normalized counts and regression coefficients listed in table 4, we calculated an Injury / control score and an AR /BK score for each validation sample. Based on the signature scores we classified each sample as C, AR, or BK.

| Group | # samples | # missed calls | # correct calls | % correct calls | comment |
| --- | --- | --- | --- | --- | --- |
| C | 100 | 41 | 59 | 59 | 16 C called AR; 25 C called BK |
| AR | 12 | 6 | 6 | 50 | 2 AR called C; 4 AR called BK |
| BK | 6 | 2 | 4 | 67 | 1 BK called C; 1 BK called AR |

## 4 DISCUSSION

Our study reports the use of a new multiplex approach to interrogate RNA isolated from urine of KTRs to identify gene signatures with clinical significance and applicability in distinguishing different causes of ongoing graft injury. The results demonstrate that a 20-gene signature identifies the presence of kidney injury vs. its absence and a 40-gene signature that may distinguish donor-reactive allo-T cell-mediated injury from BKVN. Although the incidence of TCMR in kidney grafts has been reduced by improvements in immunosuppression, it remains an important long-term risk for graft loss (30–34). Importantly, under-immunosuppression, either intentional or through non-adherence to a prescribed regimen, may lead to graft inflammation and accelerate graft loss. Overall, such a non-invasive and easily performed test might be useful in reducing the need for biopsies to differentiate causes of graft injury, allowing more personalized and timely management of immunosuppression.

In addition to the need for further independent validation, important issues that warrant further investigation include: determining the utility of the biomarker for diagnosing subclinical injury, assessing how well and how far in advance of a clinical event these gene signatures appear in the urine, and ascertaining gene expression signatures associated with other specific forms of allograft injury. Our studies provide the foundation for future work by our group among others to address these important points. This will be the focus of new investigation using serially collected urine samples from the patients studied in the validation set. In preliminary studies we have interrogated urine from a set of 19 KTRs with biopsy-proven subclinical rejection and observed an inflammatory gene expression profile in the urine of 10 patients and the absence of the profile in the other 9 (K Keslar, R Mannon, P Heeger, J Friedewald and R Fairchild, not shown). Extension of these analyses to approximately 100 patients with serially collected urine samples from CTOT-08 should be informative as to the time that the 20-gene signature appears in the urine and how it develops over time until initiation and following treatment for the subclinical and possible TCMR.

BK virus infections continue to be a major problem in kidney transplantation and BKVN is associated with graft dysfunction and failure (35–37). Reactivation of BK virus can be induced by the transplant surgery and the implementation of immunosuppression. BKVN affects more than 10% of kidney grafts and in the US may account for as many as 300 graft failures each year (38–40). The ability of the 40-gene signature to distinguish TCMR and BKVN was robust during signature development in the Training Set but was not useful during validation, due in part to the low number of samples with clinically diagnosed BKVN. An obvious limitation of the current studies is the small number of samples with TCMR and BKVN in the validation cohort, 12 and 6 samples respectively, that warrants further investigation with larger cohorts including greater numbers of these sources of kidney graft injury. At the present time we do not know if the expression of urinary transcripts specifically induced by BKVN increases with viral load in the kidney or the injury it may cause and will be another focus of future studies. Currently, urine and blood BK viral loads are monitored by qPCR; however, studies from the Suthanthiran group have reported the increase of BK virus-specific VP1 transcripts in the urine of kidney graft recipients with BKVN (41–43). Recent studies have also utilized BK specific probes to detect expression in biopsies during BKVN, including VP1, VP2, VP3 and SV40 large T antigen (44). The incorporation of probes to detect these transcripts could be a valuable addition to increase the robustness of the current 40-gene signature that distinguishes acute TCMR from BKVN and this approach could complement measurements of viremia and/or viruria in patients with suspected BKVN.

During the development of the signatures it was clear that common transcripts are upregulated in the urine during both TCMR and BKVN. These include many chemokine and proinflammatory markers and their presence is likely to be up-regulated during many inflammatory responses as has been noted previously (17–20). Whether these transcripts are restricted to inflammation in the kidney or also representative of systemic inflammation remains to be determined. Nevertheless, our analysis identified many up- and down-regulated transcripts that were specific to either acute rejection or to BKVN and these were used to develop the distinguishing 40-gene expression profile. We anticipate that a similar urine transcript-based approach could be developed to detect and distinguish other causes of kidney injury, including antibody-mediated rejection (ABMR) and calcineurin inhibitor-induced kidney injury. In the current cohort of the training and validation set patients there were no patients experiencing ABMR at the time the urine samples were collected but three patients in the cohort did develop donor-specific antibody well after this urine collection. Recent studies have identified biopsy-based gene expression profiles that are indicative of ongoing acute ABMR in kidney and heart transplants, including 6 transcripts that are associated with NK cell activation (8, 45–47). The development of a robust urine RNA-based specific for ongoing acute ABMR would not only provide a valuable non-invasive tool for detection but should be useful in monitoring the attenuation of antibody- and/or NK cell-mediated graft injury following treatment as well as the progression of antibody-mediated chronic graft injury.

As with the development of other transcript- or protein-based assays to detect the presence and cause of graft injury, a key concern with the multiplex urine RNA assay is raised by differences in the gene expression signatures observed during acute TCMR vs. the pathologic diagnosis of the graft. A clear example of such molecular and histopathologic differences is urine RNA samples from two patients with biopsy-proven acute rejection in the training set that had down-regulation of inflammatory genes and up-regulation of the same genes observed in urine RNA from the control patients. Similarly, two urine samples from recipients in the control group in the training set expressed the proinflammatory gene profile despite having no clinical evidence of graft injury. Without an available accompanying biopsy to confirm the presence or absence of graft injury it is always possible that these grafts were experiencing subclinical rejection. Within the validation test of the 20- and 40-gene signatures to detect graft injury and then distinguish acute rejection and BKVN, acute rejection was accurately detected in 50% (6 of 12) of the samples from patients with biopsy-proven rejection by the Central Pathology Core that we used for the purposes of this study to standardize histopathologic analyses. However, the 2 patient urine RNA samples procured at the time of biopsy proven rejection that the 20- gene molecular injury signature called as control/no injury had local pathology reads indicating no graft injury, raising a discrepancy in biopsy evaluation by the Core and the procurement site. The discrepancies between the molecular signatures of graft injury and the biopsy diagnosis could be interpreted as a weakness of the assay. Alternatively, as biopsy reads even by well-informed pathologists can have significant discrepancy rates (48), the molecular signature could reflect the true diagnosis: i.e. the presence or absence of graft histopathology that is not evident by microscopic evaluation or the absence of detectable transcripts indicating the injury in the urine. Similar caveats with the Edmonton molecular microscope interrogation of graft biopsy RNA vs. histopathologic evaluation have been recently raised (49).

The current results suggest the potential flexibility of the 20- and 40-gene signatures in detecting other causes of kidney graft injury that is likely to be improved with sufficient sample numbers of these and other causes of kidney graft injury and re-interrogation with the entire 796 gene target codeset to develop additional disease-specific gene expression signatures. Although the range of the current 20- and 40-gene signatures is much broader than those used in PCR-based approaches, there are likely to be additional transcript targets that increase the accuracy and sensitivity to detect and distinguish causes of graft injury. Recently, a Banff Human Organ Transplant (B-HOT) Code Set has been developed with gene sets from published data for interrogation of gene expression in graft biopsies (50). With regard the current approach, the PanCancer Immune Profiling Code Set and the B-HOT panel both have 770 probes and 403 are shared by the two code sets and 15 of the gene probes added to the panel in the current study are included in the B-HOT panel. In an effort to find additional potentially important genes expressed during TCMR, we have performed preliminary studies utilizing RNAseq analysis of urine RNA from patients with ongoing rejection and have identified up-regulated expression of many genes not present in the 796-probe code set that will be incorporated into our future probe sets to test for increased accuracy in detection of graft injury and distinction from BKVN. An important advantage of using the urine RNA vs. biopsy RNA gene signature in B-HOT and other invasive approaches is that serial samples can easily be procured and analyzed to monitor the absence vs. presence and course of kidney graft injury without the need for a biopsy. To date, we have not performed a serial interrogation of urine RNA samples from kidney transplant patients in the CTOT studies to follow progression of detected injury and the impact of recipient treatment for rejection on the expression of the signature and future studies will address these issues.

In summary, we report the initial development of a non-invasive approach to detect ongoing kidney graft injury and acute TCMR. Advantages of this approach are the ease of preparation and performance with no pre-amplification of the RNA transcripts and the ease of detection and analysis of the gene signatures. Another important strength is the flexibility of the approach in that molecular signatures for other causes of kidney graft injury are likely to lie within the 796-probe code set used in this study. As discussed, the approach to date is limited by the components of the test gene signatures. Future studies will determine if additional components can increase the specificity and sensitivity of the approach to detect T cell mediated and BKVN kidney graft injury. At the very least the interrogation of graft injury using the two gene signatures may be a valuable complementation to other molecular approaches as well as to standard of care histopathologic evaluation of the graft.

## ACKNOWLEDGEMENTS

The authors thank the participating Transplant Centers and patients for Clinical Trials in Organ Transplant Consortium CTOT-08, CTOT-10, CTOT-15, CTOT-16, and CTOT-19. This work was supported by the National Institutes of Allergy and Infectious Diseases of the National Institutes of Health under Award Numbers UO1 AI84150 (KAN), UO1 AI84146 (MMA), and U01 AI063594 (PSH) and by the Clinical Trials in Organ Transplantation Consortium NanoString Core (U01 AI063594; PSH).

## DATA AVAILABILITY STATEMENT

The data that support the findings of this study are available from the corresponding authors upon reasonable request.

## SUPPLEMENTAL MATERIALS

Additional supporting information may be found online in the Supporting Information section at the end of the article.

## Abbreviations

ABMR: antibody-mediated rejection
AUC: area under the curve
BKVN: BK virus nephropathy
CTOT: Clinical Trials in Organ Transplantation
DEG: differentially expressed genes
FSGS: focal segmented glomerular sclerosis
GEP: gene expression profiles
KTR: kidney transplant recipients
TCMR: T cell mediated rejection

