## Supplemental Table 1 for "Development of a Flexible Multiplex Urine RNA Assay for Detection and Differentiation of Kidney Allograft Injury"

Supplemental Table 1: genes added to Nanostring PanCancer Immune Profiling Panel

| **Gene** | **Full name; alias** | **Accession #** | **Category** | **Published gene set** | **Reference** |
| --- | --- | --- | --- | --- | --- |
| ADAM9 | Disintegrin and metalloproteinase domain-containing protein 9 | NM_001005845.1 | Fibrosis | Halloran IRRAT, AKI | Famulski 2012, 2013 |
| BASP1 | Brain acid soluble protein 1 | NM_006317.3 |  | Sarwal CRM | Khatri 2013 |
| CDH13 | Cadherin-13 | NM_001220488.1 | ABMR | Halloran ABMR | Sellares 2013 |
| PF4 | Platelet factor 4; CXCL4 | NM_002619.3 | Platelets |  |  |
| ACKR1 | Atypical chemokine receptor1 (Duffy blood group); DARC | NM_002036.2 | ABMR, NK | Halloran ABMR | Sellares 2013 |
| DUSP1 | Dual specificity protein phosphatase 1 | NM_004417.2 |  | Sarwal KSORT | Roedder 2014 |
| EFEMP1 | EGF-containing fibulin-like extracellular matrix protein 1 | NM_004105.3 | Fibrosis | Halloran ENDAT | Sis AJT 2009 |
| EGFR | Epidermal growth factor receptor | NM_201282.1 | Fibrosis |  | Mas 2007 |
| GBP1 | Guanylate-binding protein 1 | NM_002053.1 |  |  | Dosanjh 2013 |
| GPR171 | Probable G-protein coupled receptor 171 | NM_013308.2 | TCMR | Tcell transcript burden | Hidalgo 2012 |
| KLF4 | Krueppel-like factor 4 | NM_004235.4 | ABMR, NK | Halloran ABMR | Sellares 2013 |
| LAMC2 | Laminin subunit gamma-2 | NM_005562.2 | Fibrosis |  | Dosanjh 2013 |
| MAPK9 | Mitogen-activated protein kinase 9 | XM_005265940.1 |  | Sarwal KSORT | Roedder 2014 |
| MMP7 | matrix metallopeptidase 7 | NM_002423.3 | Fibrosis | Halloran risk score | Einecke 2010 |
| MYBL1 | V-myb avian myeloblastosis viral oncogene homolog-like 1 | XM_034274.14 | TCMR, NK | Halloran NK | Hidalgo |
| NELL2 | Protein kinase C-binding protein NELL2 | NM_006159.1 | TCMR | Tcell transcript burden | Hidalgo 2012 |
| NKG7 | Protein NKG7 | NM_005601.3 | NK cells | Sarwal CRM | Khatri 2013 |
| NKTR | NK-tumor recognition protein | NM_001012651.1 |  | Sarwal KSORT | Roedder 2014 |
| NAMPT | Nicotinamide phosphoribosyltransferase; PBEF1 | NM_005746.2 |  | Sarwal KSORT | Roedder 2014 |
| PLA1A | Phospholipase A1 member A | NM_015900.2 | ABMR, NK | Halloran ABMR | Sellares 2013 |
| ROBO4 | Roundabout homolog 4 | NM_019055.5 | ABMR, NK | Halloran ABMR | Sellares 2013 |
| SERPINA3 | Alpha-1-antichymotrypsin | NM_001085.4 | Fibrosis | Halloran IRRAT | Famulski 2012 |
| TRAC | T-cell receptor alpha chain C region; TCRA | ENST00000478163.1 | TCMR | Tcell transcript burden | Hidalgo 2012 |
| TIMP1 | Metalloproteinase inhibitor 1 | NM_003254.2 | Fibrosis |  | Anglicheau 2012 |
| TNC | Tenascin C | NM_002160.3 | Fibrosis |  | Melk 2005 |
| VCAN | chondroitin sulfate proteoglycan (versican); CSPG2 | NM_004385.3 | Fibrosis | Halloran IRRAT | Famulski 2012 |
